# KG2ML: Integrating Knowledge Graphs and Positive Unlabeled Learning for Identifying Disease-Associated Genes

**DOI:** 10.1101/2025.03.17.25323906

**Authors:** Praveen Kumar, Vincent T. Metzger, Swastika T. Purushotham, Priyansh Kedia, Cristian G. Bologa, Christophe G. Lambert, Jeremy J. Yang

## Abstract

**Background:** Biomedical knowledge graphs (KGs), such as the Data Distillery Knowledge Graph (DDKG), capture known relationships among entities (e.g., genes, diseases, proteins), providing valuable insights for research. However, these relationships are typically derived from prior studies, leaving potential unknown associations unexplored. Identifying such unknown associations, including previously unknown disease-associated genes, remains a critical challenge in bioinformatics and is crucial for advancing biomedical knowledge.

**Methods:** Traditional methods, such as linkage analysis and genome-wide association studies (GWAS), can be time-consuming and resource-intensive. This highlights the need for efficient computational approaches to identify or predict new genes using known disease-gene associations. Recently, network-based methods and KGs, enhanced by advances in machine learning (ML) frameworks, have emerged as promising tools for inferring these unexplored associations. Given the technical limitations of the Neo4j Graph Data Science (GDS) machine learning pipeline, we developed a novel machine learning pipeline called KG2ML (Knowledge Graph to Machine Learning). This pipeline utilizes our Positive and Unlabeled (PU) learning algorithm, PULSNAR (Positive Unlabeled Learning Selected Not At Random), and incorporates path-based feature extraction from ProteinGraphML.

**Results:** KG2ML was applied to 12 diseases, including Bipolar Disorder, Coronary Artery Disease, and Parkinson’s Disease, to infer disease-associated genes not explicitly recorded in DDKG. For several of these diseases, 14 out of the 15 top-ranked genes lacked prior explicit associations in the DDKG but were supported by literature and TINX (Target Importance and Novelty Explorer) evidence. Incorporating PULSNAR-imputed genes as positives enhanced XGBoost classification, demonstrating the potential of PU learning in identifying hidden gene-disease relationships.

**Conclusion:** The observed improvement in classification performance after the inclusion of PULSNAR-imputed genes as positive examples, along with the subject matter experts’ (SME) evaluations of the top 15 imputed genes for 12 diseases, suggests that PU learning can effectively uncover disease-gene associations missing from existing knowledge graphs (KGs). By integrating KG data with ML-based inference, our KG2ML pipeline provides a scalable and interpretable framework to advance biomedical research while addressing the inherent limitations of current KGs.

## Background

Genes are the fundamental units of inheritance and play a critical role in determining an individual’s susceptibility to various conditions and disorders(*MedlinePlus: Genetics, Bethesda (MD): National Library of Medicine (US)*, 2020). Mutations or genetic variations can disrupt normal biological processes, potentially leading to diseases. Identifying genes that are causally linked to specific genetic diseases is crucial for improving human health(Navlakha and Kingsford, 2010). Such knowledge provides insights into the molecular mechanisms underlying diseases, which are essential for the development of effective diagnostic and therapeutic strategies. Moreover, understanding gene-disease associations enables the identification of at-risk individuals, allowing for early interventions to reduce the likelihood of disease onset and progression(Luo *et al*., 2019; Gao *et al*., 2022; Li *et al*., 2023).

Identifying genes associated with specific diseases remains an open problem in bioinformatics(Opap and Mulder, 2017; Qumsiyeh, Showe and Yousef, 2022). Traditional approaches for disease-associated genes identification, such as linkage analysis and genome-wide association studies (GWAS), are often time-consuming and resource-intensive(Li *et al*., 2023). Consequently, the development of efficient computational methods to identify or predict novel genes using known disease-gene associations has become crucial. Network-based computational methods are widely employed to infer disease-gene associations(Gao *et al*., 2022). These networks are constructed using biological and molecular prior knowledge, capturing complex relationships between entities, such as genes, proteins, and diseases(Renaux *et al*., 2023). Knowledge graphs (KGs) further enhance this approach by integrating diverse biological networks and ontologies into a unified, comprehensive framework. KGs leverage complex semantic relationships among entities to generate valuable insights(Gao *et al*., 2022) (Renaux *et al*., 2023). Recent advancements in machine learning (ML) frameworks have expanded the application of ML algorithms to KGs for the identification and prediction of novel disease-associated genes(Mordelet and Vert, 2011; Gao *et al*., 2022; Qumsiyeh, Showe and Yousef, 2022; Li *et al*., 2023; Renaux *et al*., 2023; Gualdi, Oliva and Piñero, 2024; Xie *et al*., 2024). Applying ML methods to KGs typically involves transforming the network of entities into a feature matrix through techniques such as knowledge graph embeddings(Nelson *et al*., 2019; Gualdi, Oliva and Piñero, 2024) or path-based feature extraction methods(Himmelstein and Baranzini, 2015; Binder *et al*., 2022; Domingo-Fernández *et al*., 2022). These approaches allow the integration of heterogeneous data types and enhance the discovery of hidden patterns and associations.

In this study, we propose a machine learning pipeline called KG2ML (Knowledge Graph to Machine Learning) employing a Positive and Unlabeled (PU) learning algorithm and a path-based feature extraction method to derive features from the knowledge graph. The primary objective of this study is to identify genes potentially associated with diseases, even in the absence of explicit links between them in the knowledge graph. Since all entities and their relationships in a KG are derived from prior knowledge, the absence of a direct link between a gene and a disease does not necessarily indicate that the gene is unrelated to the disease.

This motivated the use of PU learning instead of traditional binary classification based on positive and negative examples. Specifically, we utilized PULSNAR (Positive Unlabeled Learning Selected Not At Random)(Kumar and Lambert, 2024) as our PU learning algorithm, while the method for generating feature vectors for each gene was adapted from our previous work, ProteinGraphML(Binder *et al*., 2022).

### Data Distillery Knowledge Graph (DDKG)

The Data Distillery Knowledge Graph (DDKG) has been developed as part of the Common Fund Data Ecosystem (CFDE) Data Distillery Partnership Project. This collaborative effort is led by the CFDE HuBMAP(Börner *et al*., 2025), SenNet, and Kids First teams from the University of Pittsburgh and the Children’s Hospital of Philadelphia (CHOP). Built on the Neo4j platform, the DDKG aims to integrate and distill data from multiple Common Fund data coordinating centers (DCCs), ensuring semantic interoperability to support a wide range of integrative biomedical research questions and scientific use cases, such as identifying novel relationships between biomedical entities. The DDKG schema is based on, and defined as a context of, the Unified Biomedical Knowledge Graph (UBKG)(*Unified Biomedical Knowledge Graph (UBKG)*, 2024), with schema based upon the Unified Medical Language System (UMLS)(Bodenreider, 2004) to provide a robust framework, with rigorous semantics and interoperability supported by the UMLS comprehensive metathesaurus, incorporating over 180 ontologies and standards.

### ProteinGraphML

ProteinGraphML(Binder *et al*., 2022) is a Python-based package designed to predict associations between diseases and protein-coding genes using a biomedical knowledge graph. The package employs XGBoost(Chen and Guestrin, 2016), a gradient-boosting machine learning algorithm, to estimate the likelihood of these associations. It utilizes the metapath approach to transform the complex, heterogeneous knowledge graph into a feature matrix, where rows represent proteins and columns correspond to feature vectors (either categorical or continuous variables). In the context of ProteinGraphML, a metapath is defined as a sequence of nodes and edges that form a path connecting a target protein to a disease. These metapaths capture the semantic information embedded in various path types between nodes, effectively translating the graph structure into features for machine learning models. This metapath approach allows ProteinGraphML to utilize the complex relationships within the biomedical knowledge graph, enhancing its ability to predict disease-protein associations(Binder *et al*., 2022).

### Positive Unlabeled Learning Selected Not At Random (PULSNAR)

Traditional binary classification techniques, which distinguish between positive and negative instances, are well-suited for fully labeled datasets. However, in a heterogeneous knowledge graph, the existence of a link between a disease and a gene indicates a known association, but the absence of a link does not necessarily imply a negative relationship. The gene with missing link might be positive whose association has not yet been established in studies. This leads to the issue of the lack of reliable negative examples for classification problems. This lack of reliable negative examples poses a significant challenge for traditional binary classification methods, potentially leading to biased results(Ghassemi *et al*., 2020; Kumar *et al*., 2025). To address this limitation in KG2ML, we employed PULSNAR, a Positive and Unlabeled learning technique we developed specifically for PU datasets, where reliable negative examples are missing.

Our PULSNAR package offers flexible implementation, supporting both SCAR (Selected Completely At Random) and non-SCAR scenarios, depending on the nature of the data. The package includes two main algorithms: PULSCAR (Positive Unlabeled Learning Selected Completely At Random) and PULSNAR. PULSCAR assumes that labeled positive examples are randomly selected, independent of their attributes(Elkan and Noto, 2008). In contrast, PULSNAR operates under the SNAR (Selected Not At Random) assumption, where the selection of labeled positives is dependent on their attributes(Kumar and Lambert, 2024).

PULSNAR helps identify genes that lack explicit links to a disease in the KG but may still be associated with it. For a given disease, PULSNAR estimates the proportion (ɑ) of positive genes among those without explicit links to the disease in the KG and assigns a likelihood score to each unassociated gene, indicating its probability of being a positive gene for the disease. Thus, by integrating PULSNAR, KG2ML enhances the discovery of potential disease-gene associations, particularly in cases where explicit negative examples are unavailable or unreliable.

## Materials and Methods

To apply the KG2ML pipeline to gene and disease data, we constructed a subset of the DDKG, termed *CondensedKG*, which includes all relevant node types and corresponding relationship types required for this study. A feature matrix was generated by applying a path-based approach to the nodes and edges within the CondensedKG. The subsequent subsections detail the following: (1) the methodology for creating the CondensedKG, (2) the KG2ML workflow for generating the feature matrix, (3) the limitations and challenges associated with the Neo4j Graph Data Science (GDS) library(Neo4j, 2012), (4) the identification of positive genes among those without direct links to diseases in the CondensedKG, (5) the diseases selected to evaluate the KG2ML pipeline, and (6) the validation process for imputed genes. All code necessary for constructing the CondensedKG and KG2ML pipelines are available in our GitHub repository: https://github.com/unmtransinfo/cfde-distillery.

## DDKG to CondensedKG

The DDKG integrates diverse biomedical data from multiple ontologies and DCCs, resulting in a highly complex and large-scale knowledge graph. Executing Cypher queries on such a large-scale knowledge graph to extract data for specific use cases can be resource-intensive and time-consuming. Additionally, the interpretability of the extracted data can be challenging due to its complexity. Given that our primary objective in this study is to identify gene-disease associations, we generated a subset of the DDKG, termed *CondensedKG*, to improve both data interpretability and query performance. CondensedKG comprises nodes representing genes, proteins, compounds, and diseases, along with edges that denote the relationships among these entities. Specifically, it includes 8 distinct node labels and 1,042 unique relationship types. Figure 1 illustrates the step-by-step process of generating CondensedKG from DDKG. This CondensedKG offers two key advantages: (1) it enhances data interpretability by providing a more focused and concise representation of gene-disease relationships, and (2) it significantly improves Cypher query execution times by reducing the number of nodes and edges compared to the full DDKG. Figure 2 presents the query outputs generated using DDKG and CondensedKG for the similar Neo4j Cypher query. The output produced using CondensedKG is notably more concise and interpretable compared to that using DDKG.

**Figure 1.**
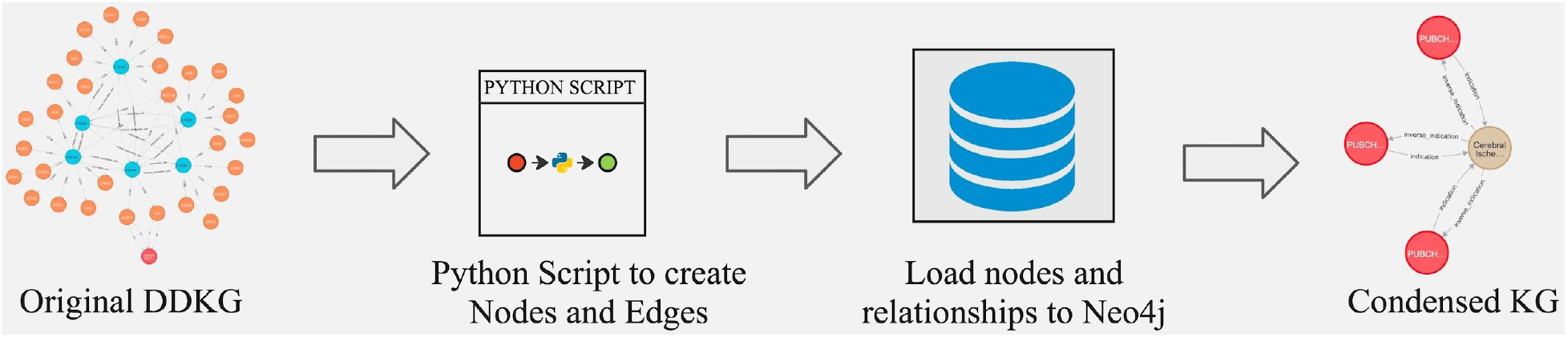
Overview of ETL workflow to generate CondensedKG from DDKG.

**Figure 2.**
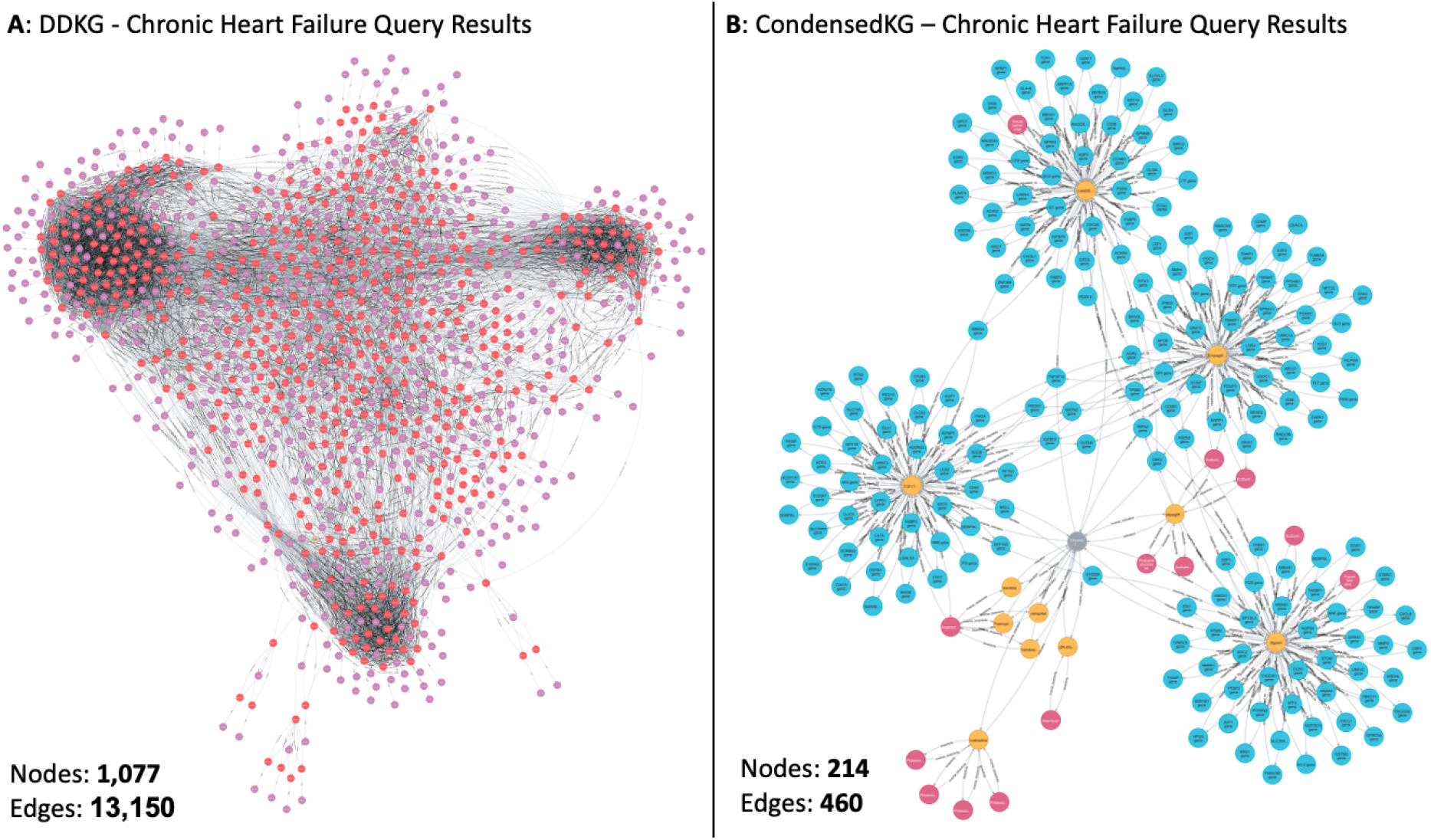
Comparison of the Chronic heart failure cypher query between DDKG and CondensedKG

### KG2ML workflow

The comprehensive workflow of the KG2ML pipeline is illustrated in Figure 3. Before applying the PULSNAR method to gene-disease data for discovering genes lacking direct links to diseases within the CondensedKG, we first identify genes with direct links to diseases (positive instances) and genes without direct links (unlabeled instances). Subsequently, we generate feature vectors for both the positive and unlabeled gene sets.

**Figure 3.**
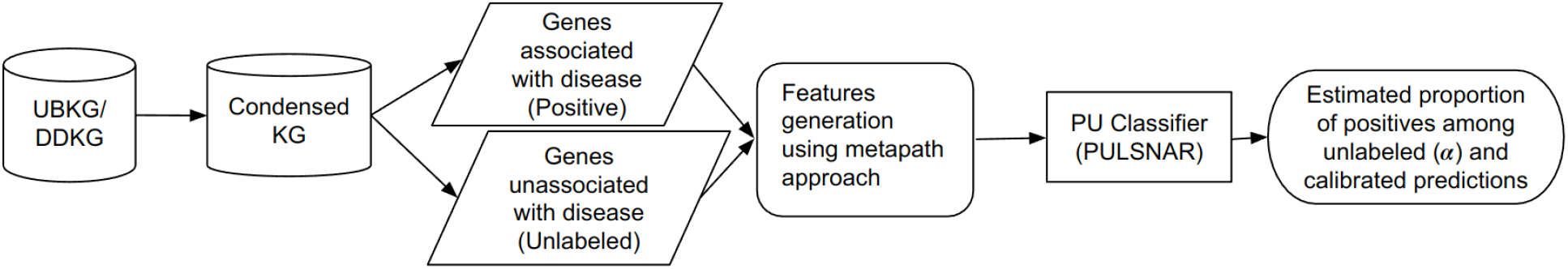
Comprehensive workflow of the KG2ML pipeline.

#### 1) Label generation

The following Cypher query was used to select all genes for inclusion in the KG2ML pipeline for Parkinson’s disease. The same query was applied to all other diseases in our study by substituting their corresponding Concept Unique Identifiers (CUIs)(*d*.*CUI* value in this query). In this query, the first MATCH statement selects all gene nodes (positive) that are directly connected to the target disease node through a compound node. The second MATCH statement selects all other gene nodes (unlabeled) that are connected to positive genes (g1:Gene) via Experimental Factor Ontology (EFO) nodes (e:EFO). For the PULSNAR method, genes identified by the first MATCH statement (g1:Gene) were labeled as class 1, while those identified by the second MATCH statement (g2:Gene) were labeled as class 0.

> *MATCH (d:Disease)-[]-(c:Compound)-[]-(g1:Gene) WHERE d*.*CUI=‘C0030567’*
>
> *MATCH (g1:Gene)-[]-(e:EFO)-[]-(g2:Gene)*
>
> *RETURN DISTINCT g1*.*CUI as positive_genes_cui, g1*.*node_label as positive_genes_label, e*.*CUI as features, c*.*CUI as features_to_remove, g2*.*CUI as unknown_genes_cui, g2*.*node_label as unknown_genes_label*

#### 2) Feature generation

All EFO nodes (e:EFO) connecting positive genes (g1:Gene) to unlabeled genes (g2:Gene) were used as features. Figure 4 visualizes the Cypher query for extracting labeled positive and unlabeled genes and the features for PU learning models.The EFO integrates components from various biological ontologies, including UBERON (anatomical terms), ChEBI (Chemical Entities of Biological Interest) chemical compounds, and the Cell Ontology. Assertions (a.k.a. semantic triples) associating genes with EFO terms are derived from several high confidence sources, with provenance available via DDKG relationship properties, encoded as source abbreviations (SABs). For each gene, if it was connected to a specific EFO node, the corresponding feature value was set to 1; otherwise, it was set to 0. This encoding resulted in a feature matrix of size n × m for running the PULSNAR models, where n represents the number of unique positive and unlabeled genes, and m denotes the number of unique features (EFO nodes). Since most genes had relatively few nonzero values (1s) in their feature vectors, the feature matrix was converted into a Compressed Sparse Row (CSR) format, significantly reducing memory usage and enhancing computational efficiency for matrix operations and storage.

**Figure 4.**
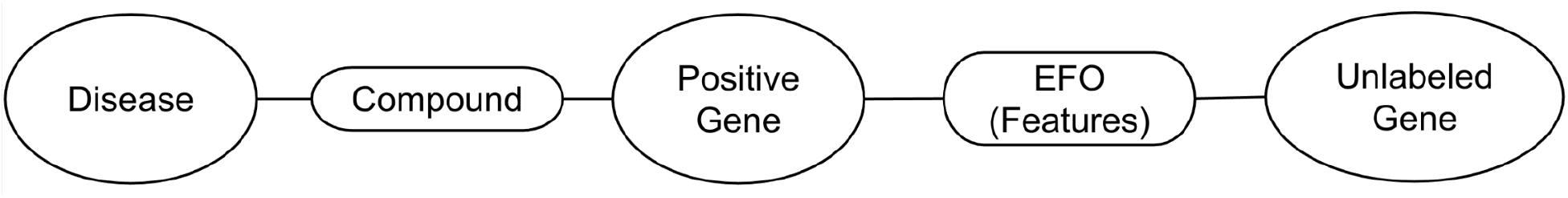
Selection of positive and unlabeled genes and extraction of associated features for the PU learning model.

### HashGNN - Neo4j Graph Data Science (GDS)

The Neo4j Graph Data Science (GDS)(Neo4j, Inc., no date) library offers machine learning pipelines for feature extraction from nodes and relationships, as well as for training supervised models to predict node properties and infer missing relationships. The library’s integration with Neo4j’s native graph storage and processing capabilities, combined with HashGNN’s theoretical ability to handle large-scale graphs efficiently, made it an attractive candidate for our biomedical knowledge graph analysis. Given the heterogeneous nature of our knowledge graph, CondensedKG, and the non-deterministic results produced by the Node2Vec node embedding algorithm — even when the randomSeed configuration parameter is specified—we exclusively utilized the HashGNN node embedding algorithm for analyzing DDKG in this study. HashGNN is particularly well-suited for embedding heterogeneous graphs and provides more deterministic results compared to algorithms like Node2Vec, especially when the randomSeed parameter is specified.

However, our experimental implementation revealed several limitations that made the HashGNN algorithm unsuitable for our KG2ML framework’s specific requirements explained in the KG2ML workflow section. While HashGNN offered efficient graph neural network operations through locality-sensitive hashing, its implementation in Neo4j GDS showed significant constraints when dealing with heterogeneous biomedical knowledge graphs. The algorithm’s performance degraded considerably when processing complex relationship types and multi-hop connections, which are characteristic of gene-disease associations. Additionally, HashGNN’s feature hashing mechanism, while efficient for homogeneous graphs, failed to adequately capture the semantic richness of different relationship types in our CondensedKG, leading to the loss of critical information about gene-disease interaction patterns.

Furthermore, beyond the HashGNN-specific limitations, Neo4j GDS presented additional challenges that affected our framework’s effectiveness. Its inability to efficiently perform batch operations for querying and processing large-scale heterogeneous knowledge graphs significantly hindered performance. Moreover, the library’s node embedding features, including HashGNN and other algorithms like FastRP and Node2Vec, lacked the flexibility required for our specific use case of disease-gene association prediction. In particular, the meta path-based approach we required for generating meaningful feature vectors that capture the complex biological relationships between genes, proteins, and diseases was not directly supported by Neo4j GDS’s embedding capabilities. These technical constraints, especially HashGNN’s limitations in handling heterogeneous graphs, led us to develop a custom path-based feature generation method that better met our specific needs for disease-gene association prediction. Therefore, this manuscript does not compare the performance of the KG2ML pipeline with the Neo4j GDS machine learning pipeline.

### Diseases used for testing the KG2ML module

Table 1 lists the diseases/conditions which were selected to evaluate the KG2ML pipeline. These well-defined diseases and conditions were specifically selected based on the following considerations: (1) availability of sufficient data, (2) biomedical importance and interest to researchers, and (3) because they are responsible for significant disease burden. Focusing on diseases with a large number of connections ensures a sufficient number of both positive and unlabeled examples, facilitating a robust evaluation of the KG2ML’s performance. Furthermore, the purposeful inclusion of a diverse range of medical conditions enabled a comprehensive evaluation of the module’s effectiveness across various diseases.

**Table 1.** List of diseases/conditions with the number of positive and unlabeled genes and features.

| Disease | UMLS_CUI | Positive genes | Unlabeled genes | Number of features |
| --- | --- | --- | --- | --- |
| Bipolar Disorder | C0005586,<br>C0236780 | 1,479 | 8,660 | 430 |
| Coronary Artery Disease | C0010068 | 1,869 | 8,286 | 505 |
| Diabetes Mellitus | C0011849,<br>C0011860,<br>C0011854 | 2,428 | 7,734 | 548 |
| Diabetic Neuropathies | C0011882 | 645 | 9,425 | 297 |
| Hodgkin Disease | C0019829 | 1,102 | 9,033 | 391 |
| Hyperaldosteronism | C0020428 | 999 | 9,140 | 365 |
| Kidney Failure | C0035078 | 2,276 | 7,886 | 524 |
| Malaria | C0024530 | 1,280 | 8,860 | 436 |
| Parkinson Disease | C0030567 | 1,964 | 8,189 | 503 |
| Pulmonary Edema | C0034063 | 1,718 | 8,425 | 467 |
| Raynaud Disease | C0034734 | 686 | 9,427 | 318 |
| Schizophrenia | C0036341 | 2,807 | 7,364 | 580 |

### Alpha estimation, identification of probable positive genes and classification performance

For all 12 diseases used in our experiment, we applied the SCAR-based algorithm available in the PULSNAR package, PULSCAR, because all positive genes for these diseases were of a single type—they were all connected to the target disease via compound nodes. Applying the non-SCAR-based method, PULSNAR, to SCAR data can lead to an overestimation of the fraction of positives among the unlabeled examples. The PULSCAR method estimated the proportion (α) of genes potentially associated with a given disease among those that lacked explicit links (unlabeled) in the CondensedKG. Using the estimated α value, PULSCAR generated calibrated probabilities for each unlabeled gene, enabling the identification of genes that were more likely to be associated with the target disease despite missing links in the CondensedKG.

In the absence of a definitive ground truth for negative genes, we employed two different modeling approaches to evaluate the performance of the XGBoost model. In the first approach, XGBoost was trained and tested using 5-fold cross-validation (CV), where all labeled positive genes were treated as class 1 and all unlabeled genes as class 0. In the second approach, XGBoost was trained and tested using 5-fold CV, but in this case, both labeled positive genes and probable positive genes identified using the PULSCAR method were treated as class 1, while the remaining unlabeled genes were treated as class 0. In both approaches, models were trained and tested using 5-fold CV for 40 iterations to estimate the 95% confidence interval (CI). This comparative analysis allowed us to assess the effectiveness of PULSCAR in distinguishing probable positive genes from truly unassociated genes.

Additionally, since all 12 datasets contained only labeled positive examples (with no explicitly labeled negative examples), we calculated recall for both models using only the labeled positive examples. Specifically, this evaluation aimed to determine whether incorporating probable positives identified by PULSCAR improved the model’s ability to recall known positive genes.

We also utilized the DEDPUL(Ivanov, 2020), TiCE(Bekker and Davis, 2018), and KM methods(Tewari, 2016) to estimate the proportion of positive genes among unlabeled genes. The publicly available implementations of the KM and TiCE methods are not optimized for large datasets, so running 40 iterations took approximately 35–40 days.

### Validation of imputed genes

To validate the genes imputed by the PULSCAR method, we selected the top 15 genes with the highest calibrated probabilities for each of the 12 diseases. The calibrated probabilities were computed as the mean values obtained from 40 independent iterations of the PULSCAR model, ensuring robustness and reliability in the probability estimates. Validation was performed by a subject matter expert (SME) through a comprehensive review of published literature and the TINX database(Metzger *et al*., 2024), a well-established resource for gene-disease associations. For each gene, the SME assessed its association with the corresponding disease based on available evidence. If a documented association was found in either the literature or the TINX database, the gene was classified as ‘Yes’ (associated). If no prior evidence of association was identified, it was classified as ‘No’ (not associated). This expert-driven validation process provided an external assessment of the PULSCAR model’s ability to identify disease-gene associations, further supporting its potential for uncovering previously unrecognized links in biomedical knowledge graphs.

## Results

Table 2 presents the α values estimated by the PULSCAR method for each of the conditions/diseases used to evaluate the KG2ML module. These α values represent the estimated proportion of genes that, despite lacking explicit links with a given disease in the knowledge graph, may potentially be associated with the condition. This suggests that a substantial fraction of unlabeled genes could have undiscovered associations with the diseases under study.

**Table 2.** α values estimated by the PULSCAR method for each disease/condition used to test the KG2ML pipeline. Recall was computed using only labeled positive genes for both models. *‘XGBoost only’*: models were trained and tested with 5-fold cross-validation using labeled positives as class 1 and unlabeled as class 0. *‘XGBoost + PULSCAR’*: models were trained and tested with 5-fold cross-validation using labeled and PULSCAR-imputed probable positives as class 1 and probable negatives as class 0.

| Disease | Estimated alpha (95% CI) | Recall (95% CI) XGBoost only | Recall (95% CI) XGBoost + PULSCAR |
| --- | --- | --- | --- |
| Bipolar Disorder | 0.3368<br>(0.3130, 0.3606) | 0.4524 (0.4502, 0.4546) | 0.6122 (0.5884, 0.6360) |
| Coronary Artery Disease | 0.3471<br>(0.3208, 0.3734) | 0.4824 (0.4803, 0.4844) | 0.6292 (0.6103, 0.6482) |
| Diabetes Mellitus | 0.2700<br>(0.2552, 0.2849) | 0.5005 (0.4992, 0.5019) | 0.5975 (0.5839, 0.6111) |
| Diabetic Neuropathies | 0.3462<br>(0.3021, 0.3903) | 0.3466 (0.3430, 0.3501) | 0.6371 (0.6088, 0.6653) |
| Hodgkin Disease | 0.4356<br>(0.4002, 0.4711) | 0.4202 (0.4174, 0.4230) | 0.6840 (0.6636, 0.7044) |
| Hyperaldosteronism | 0.4161<br>(0.3715, 0.4608) | 0.4566 (0.4538, 0.4595) | 0.6945 (0.6648, 0.7242) |
| Kidney Failure | 0.3054<br>(0.2892, 0.3217) | 0.4856 (0.4845, 0.4868) | 0.6124 (0.5901, 0.6348) |
| Malaria | 0.4273<br>(0.4148, 0.4397) | 0.4635 (0.4610, 0.4661) | 0.7177 (0.7076, 0.7277) |
| Parkinson Disease | 0.4309<br>(0.3912, 0.4708) | 0.4847 (0.4834, 0.4861) | 0.7061 (0.6793, 0.7329) |
| Pulmonary Edema | 0.3487<br>(0.3014, 0.3959) | 0.4753 (0.4735, 0.4771) | 0.6201 (0.5894, 0.6509) |
| Raynaud Disease | 0.3263<br>(0.2947, 0.3578) | 0.4269 (0.4241, 0.4297) | 0.6613 (0.6408, 0.6817) |
| Schizophrenia | 0.1514<br>(0.1235, 0.1792) | 0.5171 (0.5161, 0.5181) | 0.5483 (0.5325, 0.5641) |

In Table 2, the column labeled ‘XGBoost only’ corresponds to Model 1, which was trained and tested using 5-fold CV. In this model, all labeled positive genes were treated as class 1, and all unlabeled genes were treated as class 0. The column labeled ‘XGBoost + PULSCAR’ corresponds to Model 2, which was also trained and tested using 5-fold CV. However, in Model 2, both labeled positive genes and probable positive genes identified by the PULSCAR method were treated as class 1, while the remaining unlabeled genes were treated as class 0. The recall of Model 2 showed significant improvement over Model 1, demonstrating that incorporating probable positives identified by PULSCAR enhances the model’s ability to predict known positive genes.

Table 3 presents a comparison of alpha estimates obtained using SCAR-based methods: PULSCAR, DEDPUL, KM1, KM2, and TiCE. Since KM methods select all positive and unlabeled instances to estimate the proportion, they yielded the same alpha values across iterations. Notably, in the absence of a known true alpha for each disease, the estimated values cannot be directly validated. However, the alpha estimates from different methods suggest that some genes without direct links to the disease in existing knowledge graphs may still be associated with the disease, warranting further investigation to confirm these potential associations.

**Table 3.** Comparison of α estimated by SCAR-based methods: PULSCAR, DEDPUL, KM1, KM2, and TiCE. Estimates from PULSCAR, DEDPUL, and TiCE show the mean α and the 95% confidence interval (CI) of α values based on 40 iterations. KM estimates represent the mean α. Since the KM methods returned the same α for each iteration, 95% CI values are not shown.

| Disease | PULSCAR | DEDPUL | KM1 | KM2 | TiCE |
| --- | --- | --- | --- | --- | --- |
| Bipolar Disorder | 0.3368<br>(0.3130, 0.3606) | 0.5142<br>(0.5022, 0.5261) | 0.4547 | 0.5255 | 0.3984<br>(0.3918, 0.4050) |
| Coronary Artery Disease | 0.3471<br>(0.3208, 0.3734) | 0.4760<br>(0.4616, 0.4904) | 0.3702 | 0.4854 | 0.3989<br>(0.3917, 0.4060) |
| Bipolar Disorder | 0.3368<br>(0.3130,<br>0.3606) | 0.5142<br>(0.5022,<br>0.5261) | 0.4547 | 0.5255 | 0.3984<br>(0.3918,<br>0.4050) |
| Diabetes Mellitus | 0.2700<br>(0.2552,<br>0.2849) | 0.4633<br>(0.4513,<br>0.4753) | 0.3478 | 0.4547 | 0.3771<br>(0.3690,<br>0.3852) |
| Diabetic Neuropathies | 0.3462<br>(0.3021,<br>0.3903) | 0.5391<br>(0.5220,<br>0.5562) | 0.5193 | 0.5489 | 0.4432<br>(0.4319,<br>0.4544) |
| Hodgkin Disease | 0.4356<br>(0.4002,<br>0.4711) | 0.5239<br>(0.5145,<br>0.5332) | 0.4627 | 0.5375 | 0.4652<br>(0.4574,<br>0.4730) |
| Hyperaldosteronism | 0.4161<br>(0.3715,<br>0.4608) | 0.4844<br>(0.4772,<br>0.4916) | 0.4380 | 0.4995 | 0.4532<br>(0.4450,<br>0.4615) |
| Kidney Failure | 0.3054<br>(0.2892,<br>0.3217) | 0.4391<br>(0.4289,<br>0.4493) | 0.3702 | 0.4781 | 0.4050<br>(0.3966,<br>0.4134) |
| Malaria | 0.4273<br>(0.4148,<br>0.4397) | 0.4854<br>(0.4765,<br>0.4942) | 0.4201 | 0.4995 | 0.4537<br>(0.4482,<br>0.4592) |
| Parkinson Disease | 0.4309<br>(0.3912,<br>0.4708) | 0.4389<br>(0.4250,<br>0.4529) | 0.3809 | 0.4781 | 0.4153<br>(0.4071,<br>0.4235) |
| Pulmonary Edema | 0.3487<br>(0.3014,<br>0.3959) | 0.4923<br>(0.4823,<br>0.5023) | 0.4202 | 0.4995 | 0.3728<br>(0.3652,<br>0.3804) |
| Raynaud Disease | 0.3263<br>(0.2947,<br>0.3578) | 0.4810<br>(0.4732,<br>0.4888) | 0.4627 | 0.5128 | 0.3937<br>(0.3852,<br>0.4023) |
| Schizophrenia | 0.1514<br>(0.1235,<br>0.1792) | 0.3877<br>(0.3810,<br>0.3945) | 0.2547 | 0.4202 | 0.3185<br>(0.3135,<br>0.3235) |

Figure 5 presents the classification performance of two XGBoost-based models, highlighting the impact of incorporating PULSCAR-identified probable positive and negative genes into the classification models.

**Figure 5.**
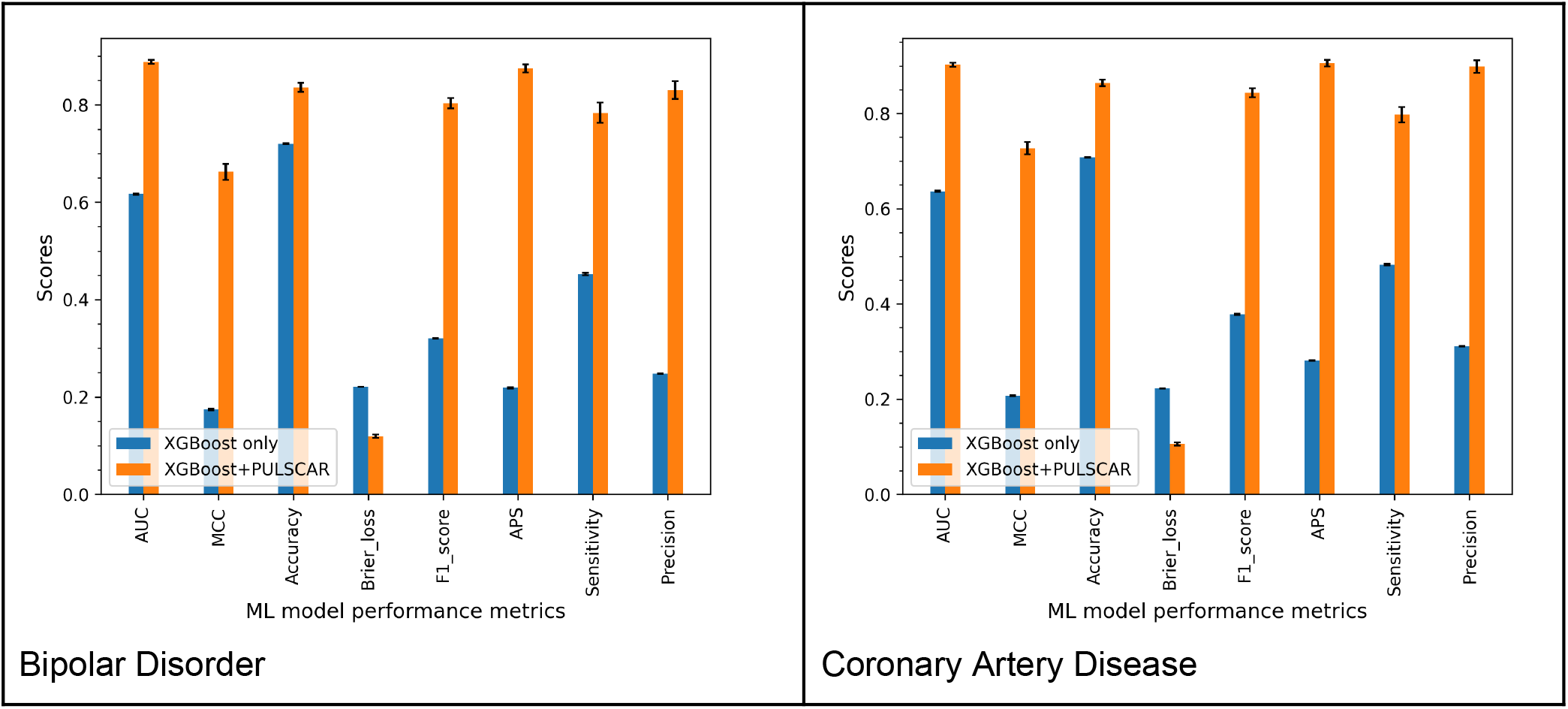
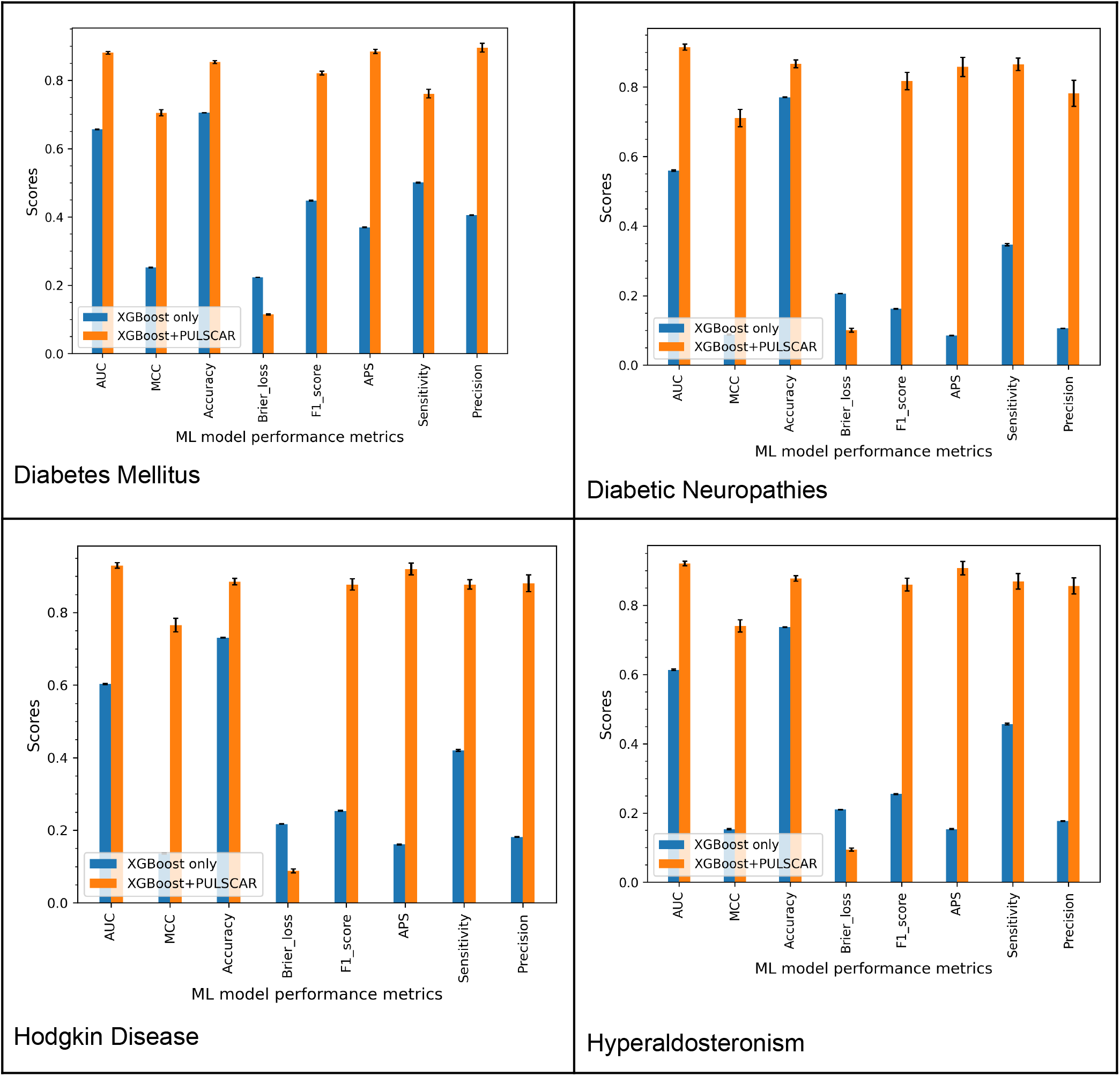
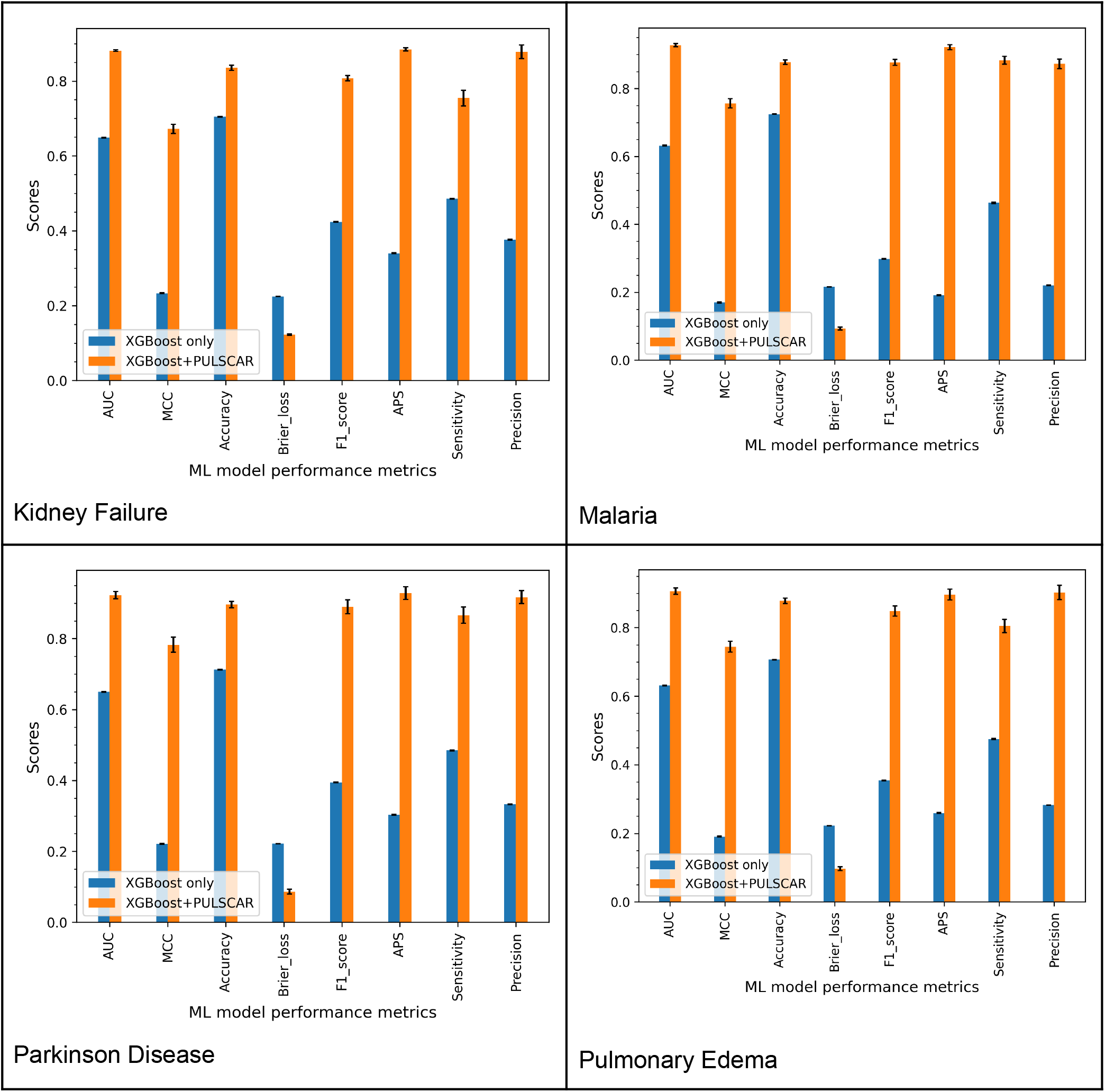
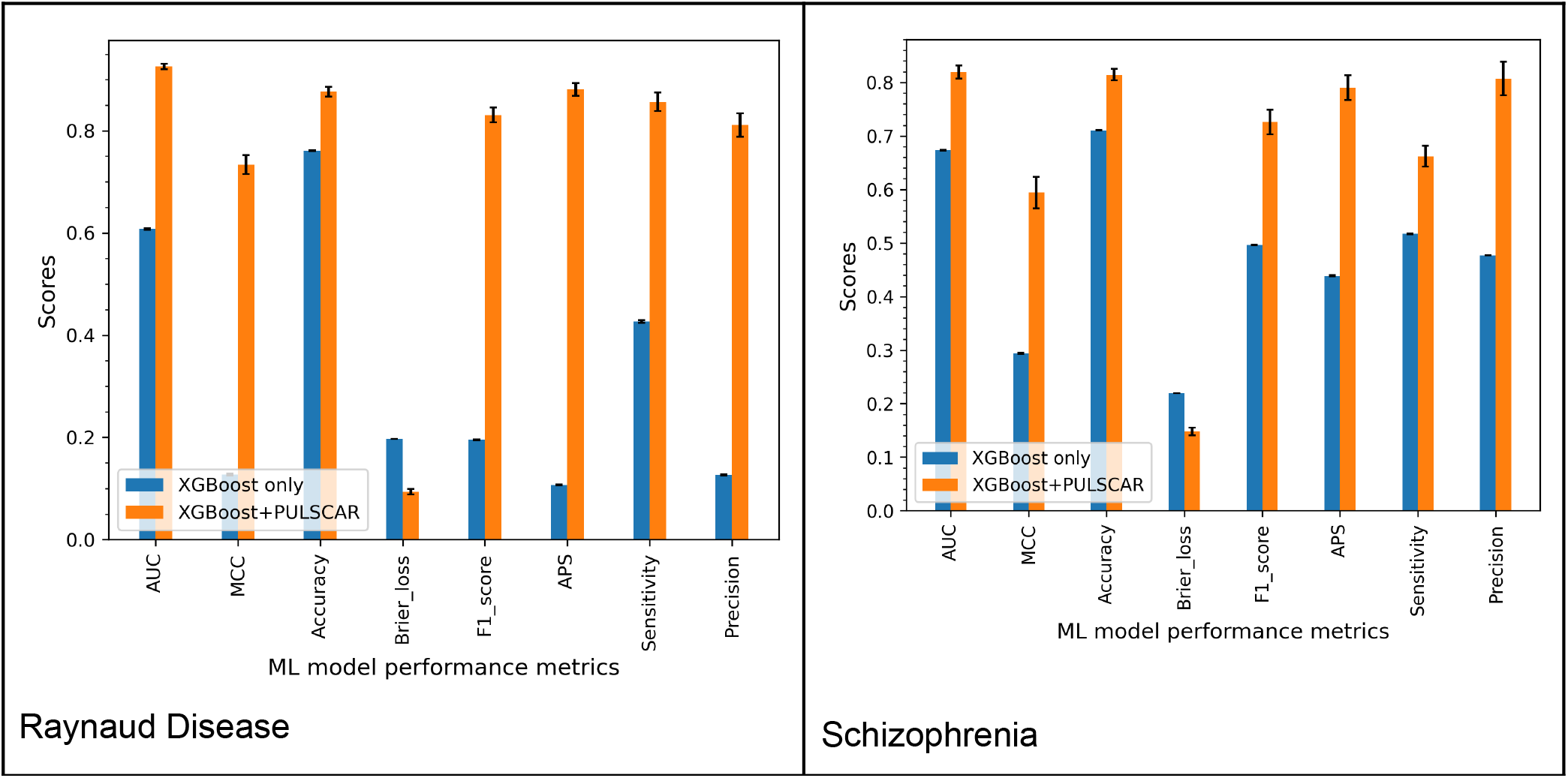
Classification performance of XGBoost models trained and tested on labeled positives as class 1 and unlabeled instances as class 0 (*‘XGBoost only’*) vs. labeled and PULSCAR-imputed probable positives as class 1 and probable negatives as class 0 (*‘XGBoost + PULSCAR’*).

Model 1 (labeled as “XGBoost only” and represented by the blue bars) was trained and tested using 5-fold cross-validation (CV). In this model, labeled positive genes were assigned to class 1, while all unlabeled genes were treated as class 0. This serves as a baseline model, operating under the assumption that all unlabeled genes are negative.

Model 2 (labeled as “XGBoost + PULSCAR” and represented by the red bars) followed the same 5-fold CV procedure. However, in this model, both labeled positive genes and probable positive genes identified by PULSCAR were assigned to class 1, while the remaining unlabeled genes were considered class 0. By incorporating the probable positive and negative genes inferred by PULSCAR, we observed a substantial improvement across all classification performance metrics across all datasets. This demonstrates the effectiveness of PULSCAR in distinguishing between positive and negative genes within the unlabeled set, thereby enhancing overall predictive accuracy.

Table 4 presents the top 15 genes with the highest calibrated probabilities of association for each of the 12 diseases analyzed in this study. Validation through a comprehensive review of existing scientific literature and the TINX database confirmed that many of these top-ranked genes are indeed associated with their respective diseases. This result highlights the effectiveness of PU learning in identifying potential disease-gene associations that are not explicitly represented in the knowledge graph.

**Table 4.**
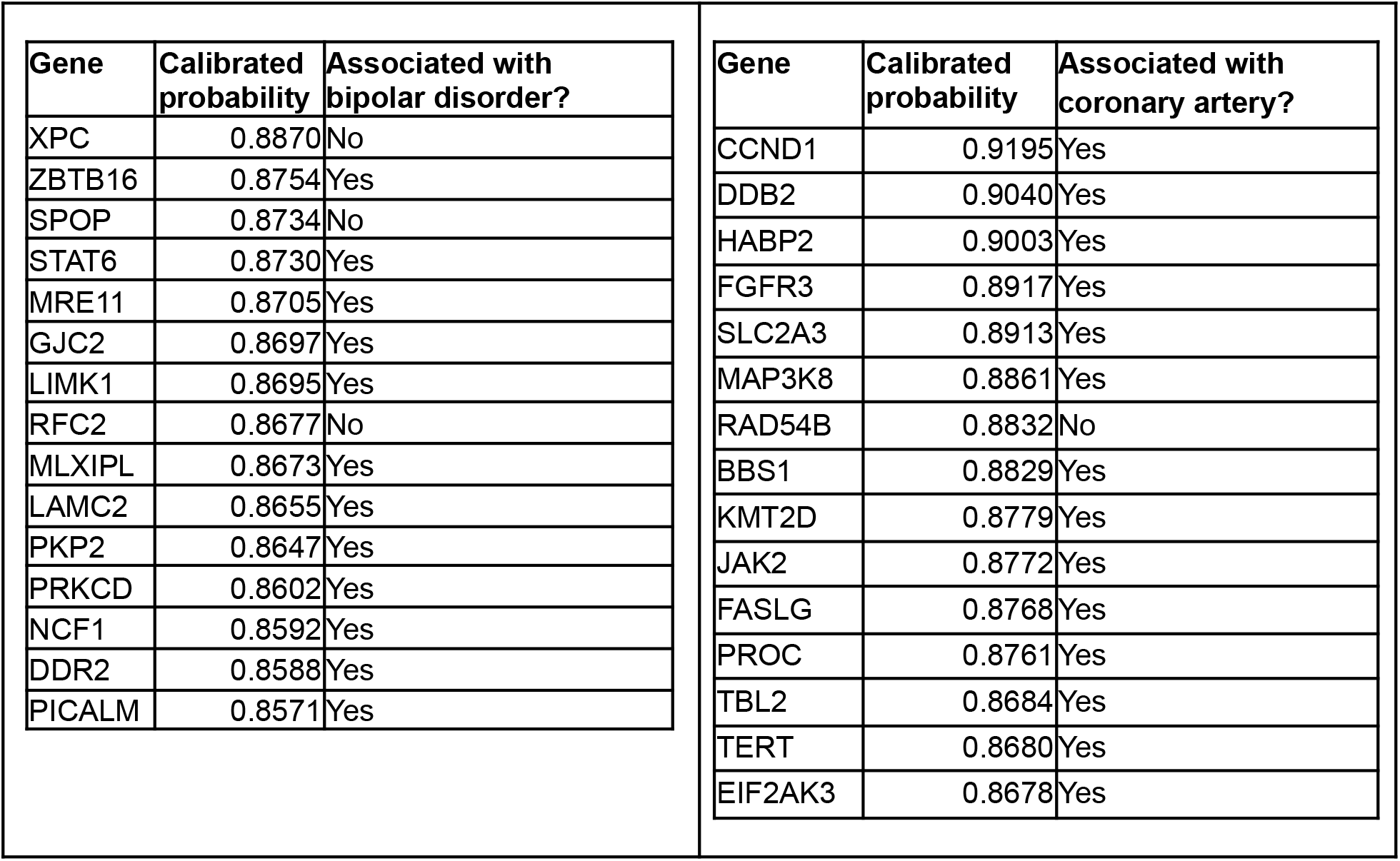
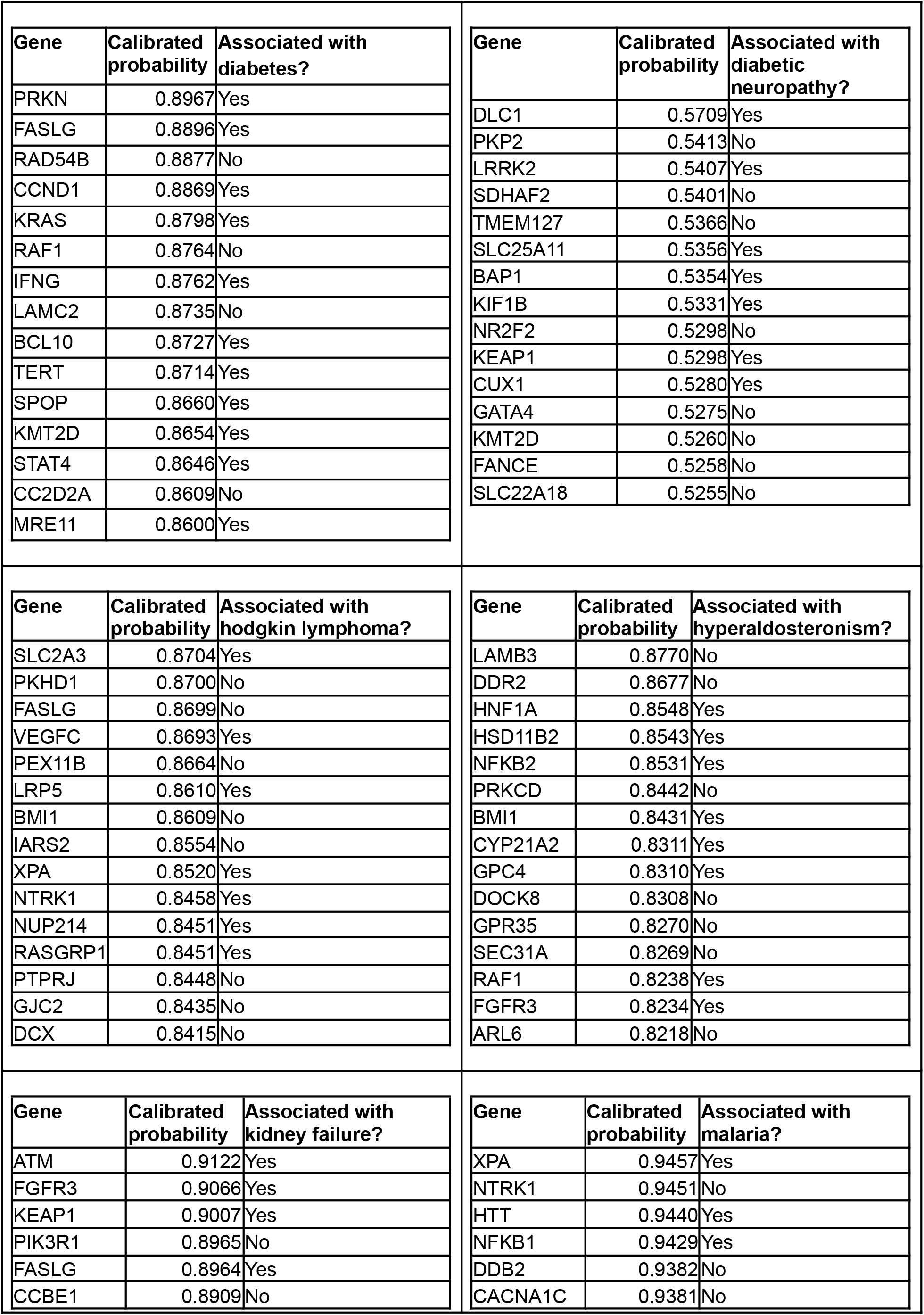
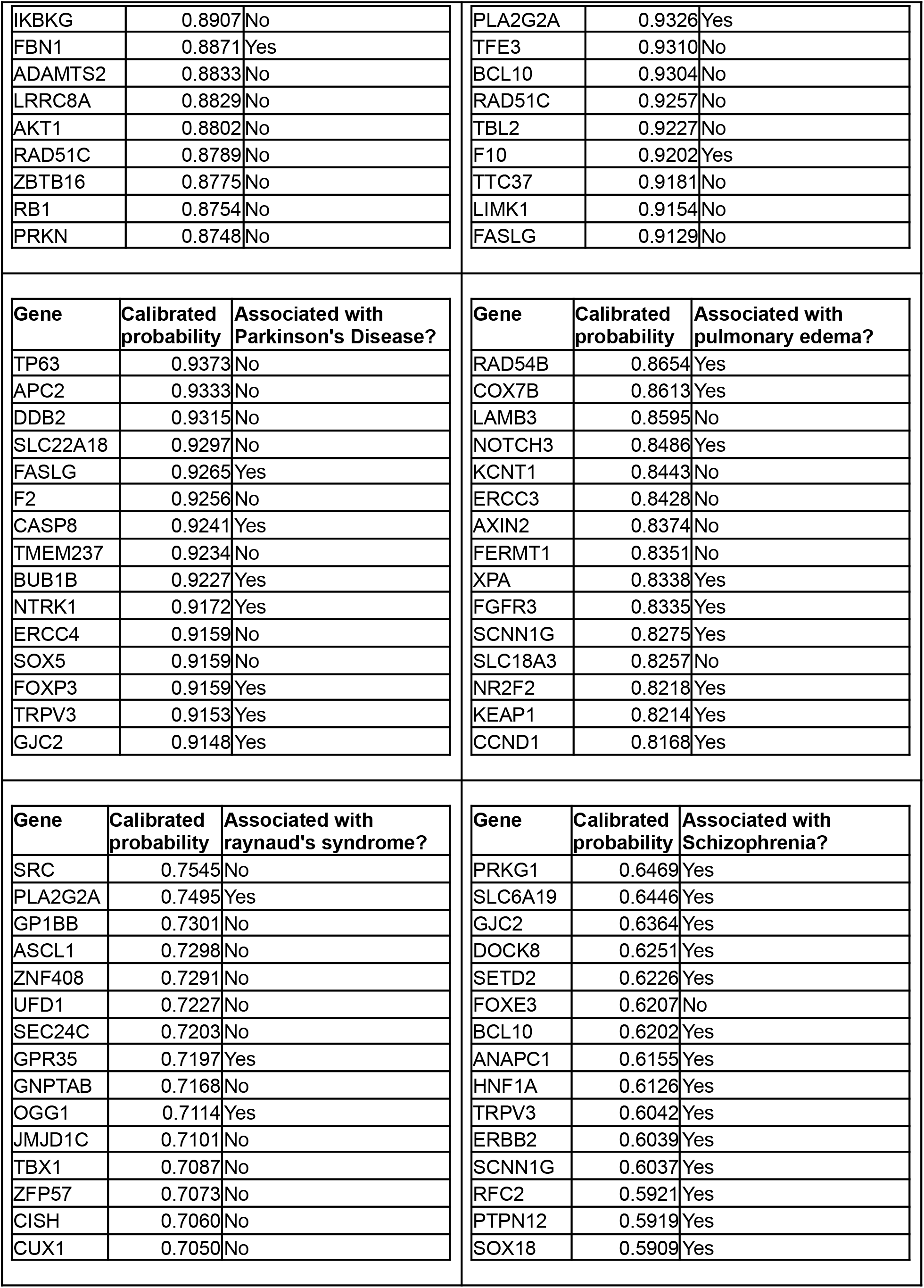
Top 15 genes with the highest calibrated probability of association for each of 12 diseases, as estimated by the PULSCAR method. Notably, several of these genes are associated with their respective diseases, even in the absence of explicit links in the knowledge graph.

## Discussion

Biomedical knowledge graphs integrate information from existing literature and databases, representing entities such as genes, diseases, and compounds as nodes and their associations as edges. However, a common limitation of these knowledge graphs is data incompleteness, as they often lack certain entities or relationships due to gaps in current knowledge(Chen *et al*., 2020). In this study, we utilized *CondensedKG*, a subset of the DDKG, which aggregates nodes and edges contributed by various DCCs. Despite its comprehensive nature, *CondensedKG*—like other knowledge graphs—remains subject to data gaps and missing associations. However, as demonstrated in our study, machine learning methods, particularly PU learning, can effectively uncover such missing associations. The α values estimated by the KG2ML pipeline for all 12 diseases in our study highlight the efficacy of PU learning in detecting genes that are not explicitly linked to diseases in the knowledge graph. However, not all imputed genes could be validated due to the novelty of these associations, as no prior studies have yet established these gene-disease relationships. The lack of validation for certain gene-disease pairs does not necessarily indicate that the associations identified by the PU learning method are incorrect; rather, it suggests the potential for novel discoveries that warrant further investigation.

Knowledge graph embedding algorithms map entities and relationships from a knowledge graph into a continuous vector space while preserving the graph’s semantic and structural properties(Dai *et al*., 2020). These embeddings enable efficient computation and facilitate machine learning tasks such as link prediction and entity classification. However, as outlined in the subsection “HashGNN - Neo4j Graph Data Science (GDS),” Neo4j’s graph embedding algorithms can exhibit non-deterministic behavior despite setting a random seed, which poses challenges for reproducibility. To address this limitation, we adopted a path-based embedding technique from our previous work, ProteinGraphML, ensuring more consistent and interpretable feature vector representations. This approach not only enhanced reproducibility but also provided embeddings better suited for the downstream PU learning method, PULSCAR.

Across all datasets, Model 2 consistently outperformed Model 1 across all evaluation metrics (Figure 5 and Table 2). This improvement suggests that the application of PU learning effectively differentiated probable positive and negative examples within the unlabeled set, thereby refining the dataset. The improved data quality enabled the model to learn more precise decision boundaries, resulting in superior classification performance compared to the baseline model (Model 1), which treated all unlabeled examples as negative instances (class 0). This finding is consistent with prior studies demonstrating that reducing noise in the labels improves model performance(Ding *et al*., 2022). A review of PU learning in bioinformatics and computational biology(Li *et al*., 2021) also found that studies reported performance improvements when using PU learning methods. The significant improvement in recall, as shown in Table 2, further supports the accuracy of our PU method, PULSCAR, in identifying probable positives within the unlabeled set. Since the recall was calculated using only labeled positive examples, the results suggest that the probable positives identified by PULSCAR were predominantly true positives rather than false positives. Our findings highlight the potential of PU learning to improve gene-disease association predictions, making it a valuable computational approach for advancing biomedical research.

### Limitations

To validate the genes identified by the KG2ML module, we referenced published scientific literature along with the TINX database. If a gene-disease association was neither documented in prior studies nor available in TINX, we could not confirm its validity based on the existing knowledge. Consequently, our findings should be interpreted as methodological advancements in identifying previously unrecognized disease-associated genes, rather than as definitive associations.

### Future work

In this study, we were unable to utilize the Neo4j Graph Data Science module due to its technical constraints, as outlined in the subsection “HashGNN - Neo4j Graph Data Science (GDS).” In future research, we aim to integrate a deterministic node embedding algorithm of Neo4j with our KG2ML pipeline. This investigation will evaluate the effectiveness of GDS-derived embeddings in enhancing PU learning performance for identifying disease-gene associations.

## Conclusions

Like other biomedical knowledge graphs, *CondensedKG*, despite being a subset of the comprehensive DDKG, lacks certain associations among existing entities such as genes, diseases, compounds, and proteins. Since all gene-disease associations in *CondensedKG* are derived from prior studies and established databases, genes explicitly linked to a disease can be considered positive genes for that disease. However, genes lacking such associations in the knowledge graph cannot be assumed to be negative genes, as their true relationships may simply be undiscovered. This characteristic makes PU learning particularly suitable for analyzing such data, as it is specifically designed to identify unknown associations in the absence of confirmed negative examples. The α values estimated by PULSCAR suggest that numerous missing gene-disease associations exist in *CondensedKG*, highlighting the potential for novel discoveries. However, these predicted associations require experimental validation to confirm their biological relevance. Manual validation of imputed genes by domain experts further demonstrates the potential of PU learning as a computational framework for advancing biomedical research. By integrating knowledge graph analysis with PU learning, the KG2ML pipeline presents a methodological advancement, providing a robust and scalable framework for uncovering novel gene-disease associations.

## Data Availability

All data produced are available online at https://github.com/unmtransinfo/cfde-distillery.

## Declarations

### Ethics approval and consent to participate

Not applicable.

### Consent for publication

Not applicable.

### Availability of data and material

The original accession numbers, names of the databases and data repositories, and permanent web links (URLs) corresponding to the datasets supporting the findings of this study, plus all source code as needed for reproducibility, are available via GitHub repository https://github.com/unmtransinfo/cfde-distillery/, specifically the README. Data files are all standard formats, with rigorous metadata including community-standard identifiers, such as PubChem chemical IDs, UniProt protein IDs, Ensembl gene IDs, UMLS disease Concept Unique Identifiers (CUIs), Pharos target IDs, and DrugCentral drug IDs.

### Author contributions

PKumar, VM, and JY conceived the study. PKumar, VM, SP, and PKedia developed the ETL workflows, KG2ML analytics, ML methodology, and biomedical use cases. CB and CL advised on computational and biomedical issues. JY supervised the research and technical contributors. PKumar, VM, and JY wrote the manuscript. All authors reviewed and approved the manuscript.

## Acknowledgments

We gratefully acknowledge UBKG/DDKG leaders, designers, and developers Jonathan Silverstein and Alan Simmons, for informative discussions and assistance.

## References

Bekker, J. and Davis, J. (2018) ‘Estimating the class prior in positive and unlabeled data through decision tree induction’, Proceedings of the … AAAI Conference on Artificial Intelligence. AAAI Conference on Artificial Intelligence, 32(1). Available at: 10.1609/aaai.v32i1.11715.

Binder, J. et al. (2022) ‘Machine learning prediction and tau-based screening identifies potential Alzheimer’s disease genes relevant to immunity’, Communications biology, 5(1), p. 125.

Bodenreider, O. (2004) ‘The Unified Medical Language System (UMLS): integrating biomedical terminology’, Nucleic acids research, 32(Database issue), pp. D267–70.

Börner, K. et al. (2025) ‘Human BioMolecular Atlas Program (HuBMAP): 3D Human Reference Atlas construction and usage’, Nature methods, pp. 1–16.

Chen, T. and Guestrin, C. (2016) ‘XGBoost’, in Proceedings of the 22nd ACM SIGKDD International Conference on Knowledge Discovery and Data Mining. KDD ‘16: The 22nd ACM SIGKDD International Conference on Knowledge Discovery and Data Mining, New York, NY, USA: ACM. Available at: 10.1145/2939672.2939785.

Chen, Z. et al. (2020) ‘Knowledge Graph Completion: A Review’, IEEE access: practical innovations, open solutions, 8, pp. 192435–192456.

Dai, Y. et al. (2020) ‘A survey on knowledge graph embedding: Approaches, applications and benchmarks’, Electronics, 9(5), p. 750.

Ding, C. et al. (2022) ‘Impact of Label Noise on the Learning Based Models for a Binary Classification of Physiological Signal’, Sensors (Basel, Switzerland), 22(19). Available at: 10.3390/s22197166.

Domingo-Fernández, D. et al. (2022) ‘Causal reasoning over knowledge graphs leveraging drug-perturbed and disease-specific transcriptomic signatures for drug discovery’, PLoS computational biology, 18(2), p. e1009909.

Elkan, C. and Noto, K. (2008) ‘Learning classifiers from only positive and unlabeled data’, in Proceedings of the 14th ACM SIGKDD international conference on Knowledge discovery and data mining. KDD08: The 14th ACM SIGKDD International Conference on Knowledge Discovery and Data Mining, New York, NY, USA: ACM.

Gao, Z. et al. (2022) ‘A knowledge graph-based disease-gene prediction system using multi-relational graph convolution networks’, AMIA … Annual Symposium proceedings. AMIA Symposium, 2022, pp. 468–476.

Ghassemi, M. et al. (2020) ‘A Review of Challenges and Opportunities in Machine Learning for Health’, AMIA Joint Summits on Translational Science proceedings. AMIA Joint Summits on Translational Science, 2020, pp. 191–200.

Gualdi, F., Oliva, B. and Piñero, J. (2024) ‘Predicting gene disease associations with knowledge graph embeddings for diseases with curtailed information’, NAR genomics and bioinformatics, 6(2), p. lqae049.

Himmelstein, D.S. and Baranzini, S.E. (2015) ‘Heterogeneous Network Edge Prediction: A Data Integration Approach to Prioritize Disease-Associated Genes’, PLoS computational biology, 11(7), p. e1004259.

Ivanov, D. (2020) ‘DEDPUL: Difference-of-estimated-densities-based positive-unlabeled learning’, in 2020 19th IEEE International Conference on Machine Learning and Applications (ICMLA). 2020 19th IEEE International Conference on Machine Learning and Applications (ICMLA), IEEE. Available at: 10.1109/icmla51294.2020.00128.

Kumar, P. et al. (2025) ‘Detecting Opioid Use Disorder in Health Claims Data With Positive Unlabeled Learning’, IEEE journal of biomedical and health informatics, 29(2), pp. 750–757.

Kumar, P. and Lambert, C.G. (2024) ‘Positive Unlabeled Learning Selected Not At Random (PULSNAR): class proportion estimation without the selected completely at random assumption’, PeerJ. Computer science, 10, p. e2451.

Li, F. et al. (2021) ‘Positive-unlabeled learning in bioinformatics and computational biology: a brief review’, Briefings in Bioinformatics, 23(1), p. bbab461.

Li, Y. et al. (2023) ‘End-to-end interpretable disease-gene association prediction’, Briefings in bioinformatics, 24(3). Available at: 10.1093/bib/bbad118.

Luo, P. et al. (2019) ‘Identifying Disease-Gene Associations With Graph-Regularized Manifold Learning’, Frontiers in genetics, 10, p. 270.

MedlinePlus: Genetics, Bethesda (MD): National Library of Medicine (US) (2020) MedlinePlus: Genetics. Available at: https://medlineplus.gov/genetics (Accessed: 5 January 2025).

Metzger, V.T. et al. (2024) ‘TIN-X version 3: update with expanded dataset and modernized architecture for enhanced illumination of understudied targets’, PeerJ, 12, p. e17470.

Mordelet, F. and Vert, J.-P. (2011) ‘ProDiGe: Prioritization Of Disease Genes with multitask machine learning from positive and unlabeled examples’, BMC bioinformatics, 12, p. 389.

Navlakha, S. and Kingsford, C. (2010) ‘The power of protein interaction networks for associating genes with diseases’, Bioinformatics (Oxford, England), 26(8), pp. 1057–1063.

Nelson, W. et al. (2019) ‘To Embed or Not: Network Embedding as a Paradigm in Computational Biology’, Frontiers in genetics, 10, p. 381.

Neo4j (2012) Neo4j - The World’s Leading Graph Database. Available at: http://neo4j.org/ (Accessed: 5 January 2025).

Neo4j, Inc. (no date) Neo4j Graph Platform, Neo4j. Available at: https://neo4j.com (Accessed: 21 December 2020).

Opap, K. and Mulder, N. (2017) ‘Recent advances in predicting gene-disease associations’, F1000Research, 6, p. 578.

Qumsiyeh, E., Showe, L. and Yousef, M. (2022) ‘GediNET for discovering gene associations across diseases using knowledge based machine learning approach’, Scientific reports, 12(1), p. 19955.

Renaux, A. et al. (2023) ‘A knowledge graph approach to predict and interpret disease-causing gene interactions’, BMC bioinformatics, 24(1), p. 324.

Tewari, H.R.C.S. (2016) ‘Mixture proportion estimation via kernel embeddings distributions’, in Proceedings of The 33rd International Conference on Machine Learning, PMLR 48. The 33rd International Conference on Machine Learning.

Unified Biomedical Knowledge Graph (UBKG) (2024) The Unified Biomedical Knowledge Graph (UBKG) is a knowledge graph infrastructure that represents a set of interrelated concepts from biomedical ontologies and vocabularies. Available at: https://ubkg.docs.xconsortia.org/ (Accessed: 5 January 2025).

Xie, J. et al. (2024) ‘Predicting disease-gene associations through self-supervised mutual infomax graph convolution network’, Computers in biology and medicine, 170, p. 108048.

